# Assessing dengue forecasting methods: A comparative study of statistical models and machine learning techniques in Rio de Janeiro, Brazil

**DOI:** 10.1101/2024.06.12.24308827

**Authors:** Xiang Chen, Paula Moraga

## Abstract

**Background:** Dengue is a mosquito-borne viral disease that poses a significant public health threat in tropical and subtropical regions worldwide. Accurate forecasting of dengue outbreaks is crucial for effective public health planning and intervention. This study aims to assess the predictive performance and computational efficiency of a number of statistical models and machine learning techniques for dengue forecasting, both with and without the inclusion of climate factors, to inform the design of dengue surveillance systems.

**Methods:** The study considers dengue cases in Rio de Janeiro, Brazil, as well as climate factors known to affect disease transmission. Employing a dynamic window approach, various statistical methods and machine learning techniques were used to generate weekly forecasts at several time horizons. Error measures, uncertainty intervals, and computational efficiency obtained with each method were compared. Statistical models considered were Autoregressive (AR), Moving Average (MA), Autoregressive Integrated Moving Average (ARIMA), and Exponential Smoothing State Space Model (ETS). Additionally, models incorporating temperature and humidity as covariates, such as Vector Autoregression (VAR) and Seasonal ARIMAX (SARIMAX), were employed. Machine learning techniques evaluated were Random Forest, XGBoost, Support Vector Machine (SVM), Long Short-Term Memory (LSTM) networks, and Prophet. Ensemble approaches that integrated the top performing methods were also considered. The evaluated methods also incorporated lagged climatic variables to account for delayed effects.

**Results:** Among the statistical models, ARIMA demonstrated the best performance using only historical case data, while SARIMAX significantly improved predictive accuracy by incorporating climate covariates. In general, the LSTM model, particularly when combined with climate covariates, proved to be the most accurate machine learning model, despite being slower to train and predict. For long-term forecasts, Prophet with climate covariates was the most effective. Ensemble models, such as the combination of LSTM and ARIMA, showed substantial improvements over individual models.

**Conclusion:** This study demonstrates the strengths and limitations of various methods for dengue forecasting across multiple timeframes. It highlights the best-performing statistical and machine learning methods, including their computational efficiency, underscoring the significance of machine learning techniques and the integration of climate covariates to improve forecasts. These findings offer valuable insights for public health officials, facilitating the development of dengue surveillance systems for more accurate forecasting and timely allocation of resources to mitigate dengue outbreaks.

**Author summary:** Dengue is a mosquito-borne viral disease that poses a significant public health threat in tropical and subtropical regions worldwide. Accurate forecasting of dengue can significantly aid in public health planning and response. In this study, we compared the performance of various statistical models and machine learning techniques to predict dengue cases across several timeframes. In the evaluation, we used historical dengue case data in Rio de Janeiro, Brazil, as well as climate factors such as temperature and humidity known to affect transmission. Methods considered included traditional statistical models like ARIMA and SARIMAX, and advanced machine learning approaches like Random Forest, XGBoost, SVM, LSTM, and Prophet. We found that integrating climate data significantly improved the accuracy of forecasts. Specifically, the LSTM model combined with climate covariates provided the most accurate predictions overall, while Prophet was particularly effective for long-term forecasts. Additionally, ensemble approaches that combined multiple models outperformed individual models. This work demonstrates the potential of machine learning techniques to provide timely and accurate predictions, and emphasizes the importance of climate data in dengue forecasting. The study aims to support public health officials in developing dengue surveillance systems to enable informed decision-making for mitigating the impact of dengue outbreaks.

## Introduction

Dengue is a viral infection transmitted through the bites of infected female *Aedes* mosquitoes. The main vector for dengue, the *Aedes aegypti* mosquito, thrives in high temperature and humidity, conditions that also promote viral replication. The dengue virus exists in four distinct serotypes, namely, DENV-1, DENV-2, DENV-3, and DENV-4. Infection with any serotype can result in dengue fever, manifesting from mild febrile illness to severe forms such as dengue hemorrhagic fever or dengue shock syndrome, which can be fatal.

Dengue represents a significant public health concern in tropical and subtropical regions in the world and is now established as endemic in over hundred of countries across Africa, the Americas, the Eastern Mediterranean, South-East Asia, and the Western Pacific [1]. Moreover, dengue cases are projected to increase and expand into new territories in the coming years as climate change alters epidemiological patterns [2]. Dengue not only affects population health but also places a significant economic burden on a global scale.

Currently, there is no universal treatment for dengue. However, various prevention strategies are available, including personal protection, chemical control, and environmental management of *Aedes* mosquitoes [3]. Enhanced surveillance is also crucial for early detection and response to dengue outbreaks allowing public health authorities to respond promptly and implement control measures to allocate resources in areas of higher risk and prevent further spread.

Forecasting dengue incidence involves various methodologies that range from traditional statistical models to advanced machine learning techniques. Each method offers distinct advantages and limitations in terms of accuracy, complexity, and applicability. Traditional statistical methods, such as ARIMA (Autoregressive Integrated Moving Average) and seasonal decomposition, have been widely used for infectious disease forecasting. These models are particularly effective at capturing linear relationships and seasonal patterns in time-series data. However, they often require stationary data and may struggle to handle non-linear interactions or abrupt changes in trends. For instance, [4] developed seasonal ARIMA models for forecasting dengue in northeastern Thailand. Their models provided detailed insights into the timing and intensity of outbreaks, proving practical for public health planning. [5] utilized ARIMA models to predict dengue incidence in Rio de Janeiro, Brazil. Their findings suggested that simple models could be effective, particularly when predicting short-term outcomes, highlighting the value of using lagged cases as predictors to enhance forecast accuracy. ARIMA models were also utilized by [6] to analyze dengue incidence in Recife and Goiania, Brazil, emphasizing the importance of both trend and seasonality. The study highlighted the challenges of forecasting during the Zika virus co-circulation, which interfered with accurate dengue case predictions. [7] combined various statistical models to predict peak dengue transmission in Iquitos, Peru. This strategy proved superior in estimating the peak heights and total case counts, illustrating the advantages of combining diverse methodologies.

In recent years, machine learning models have gained prominence due to their ability to learn complex patterns from data without explicit programming. Techniques like Random Forests, Support Vector Machines (SVM), and neural networks, including Long Short-Term Memory (LSTM) networks, offer powerful alternatives to traditional models. These methods can capture complex non-linear interactions and are generally more flexible in handling various data types and structures. Nevertheless, machine learning models often require large datasets for training and can be opaque, making interpretation challenging. [8] developed and compared various machine learning algorithms in Brazilian cities, showing that Random Forests, when optimized with local data, yielded the lowest prediction errors. [9] applied multiple models including LSTM and support vector regression in Kerala, India, with LSTM providing the most accurate predictions for dengue prevalence, illustrating the model’s superior capability to capture complex patterns in disease spread. [10] introduced a hybrid deep learning architecture combining convolutional and recurrent neural networks for forecasting dengue incidence. This novel approach proved highly effective, offering significant improvements in forecasting accuracy over other deep learning models.

Incorporating climate and environmental information has been shown to enhance both statistical and machine learning models. Temperature, rainfall, and humidity have been linked to the breeding and survival rates of mosquitoes, thereby affecting dengue transmission rates [11]. Models that integrate these environmental factors tend to perform better in predicting dengue incidence over longer time horizons, reflecting the complex interplay between the pathogen, host, and environment [12–15]. For instance, [12] improved dengue surveillance in Guadeloupe, French West Indies, by integrating climatic variables into SARIMA (Seasonal ARIMA) models. This approach not only forecasted outbreaks with higher accuracy but also demonstrated the specific impacts of temperature and humidity on dengue incidence. [16] analyzed dengue fever outbreaks in Selangor, Malaysia, using several machine learning techniques with climate variables as predictors. The study found that SVM (linear kernel) exhibited the best overall performance, particularly in terms of specificity and precision. [17] explored the efficiency of Random Forests and artificial neural networks to forecast dengue in Colombia, demonstrating that Random Forests was superior for short-to medium-term dengue predictions at national and departmental levels, benefiting from the integration of socio-demographic and environmental predictors.

In this paper, we evaluate a number of statistical models and machine learning techniques for dengue forecasting. The aim is to determine which methods provide superior predictive performance and computational efficiency, which could inform the design of dengue surveillance systems. Several studies have compared the performance of various dengue prediction methods [8, 16–21]. However, these studies have several gaps which we aim to address in our evaluation. First, unlike previous studies which often focus on monthly predictions, we consider forecasts at a weekly resolution. This finer granularity provides a more precise and actionable scale for public health decision-making. In addition, in contrast to other works that typically use a fixed window approach to assess performance, we adopt a moving window strategy where models are continuously trained with newly acquired data. This approach reflects what would happen in real-time scenarios where data is continuously updated. Moreover, this approach allows us to capture the evolving patterns and trends of dengue, enhancing models’ robustness and accuracy over time.

Understanding the uncertainty associated with forecasts is critical for public health decision-making. Our study computes uncertainty intervals and coverage probabilities, providing a more comprehensive understanding of the predictions’ reliability. Many other studies only report point predictions, which do not convey the inherent uncertainties in the forecasts. In addition to single models, our research also compares ensemble approaches that combine multiple forecasting methods that leverage the unique strengths of individual models to enhance predictive accuracy. The consideration of ensembles highlight the potential of these approaches for dengue forecasting.

Additionally, we assess the models’ performance across several forecast horizons, from short-term (1-4 weeks) to long-term (8-12 weeks) predictions. In contrast to other studies that often focus on a single forecast horizon, this comprehensive evaluation allows us to understand the models’ strengths and limitations across different time scales, providing a more detailed picture of their forecasting capabilities. Finally, we report the computational time required to run each forecasting method. This aspect is particularly important in settings with limited resources, where the time and cost associated with running complex models can be prohibitive.

In our evaluation, we utilize weekly cases in Rio de Janeiro, Brazil, a region that faces frequent dengue epidemics. Dengue cases were obtained from InfoDengue [22], a system that partially automates the collection, organization, and analysis of climate and epidemiological data of dengue and other arboviruses in municipalities across Brazil.

The methods considered in the analysis include statistical and machine learning methods known for their demonstrated effectiveness in time series forecasting applied to epidemiology. Specifically, we utilize the statistical methods Autoregressive (AR), Moving-Average (MA), ARIMA [23, 24], and Exponential Smoothing (ETS) [25], known for their robustness in analyzing time-dependent patterns. We also consider Seasonal ARIMA with eXogenous variables (SARIMAX) [24] and Vector Autoregression (VAR) [23, 26] models, which allow us to integrate external variables and assess their impact alongside historical disease trends. Additionally, we explore machine learning techniques like Random Forest [27], XGBoost [28], SVM [29, 30], LSTM [31, 32], and Prophet [33]. These methods are selected for their advanced capabilities in modeling complex interactions and non-linear relationships [34, 35], applicable both with and without climate covariates. Finally, we consider ensemble approaches that combine statistical and machine learning approaches.

The rest of the paper is organized as follows. In Section Dengue and climate data in Rio de Janeiro, Brazil, we describe the study region and the dengue and climate data utilized for the comparison of the methods. Section Methodology covers the statistical methods and machine learning techniques employed, as well as the evaluation metrics for comparison. Section Results presents the results, categorizing them into statistical models, machine learning techniques, and ensemble approaches, both with and without covariates. This illustrates the effectiveness of each method along with their computational efficiency. The paper concludes with a discussion of the findings and their implications for dengue surveillance.

### Dengue and climate data in Rio de Janeiro, Brazil

Rio de Janeiro, situated in southeastern Brazil, is a vast urban area known for its high population density and subtropical climate (Figure 1). With a population exceeding 16 million [36], the city is particularly susceptible to dengue fever, largely because of the conducive environment for the proliferation of *Aedes aegypti* mosquitoes. The warm, humid climate in the area provides an optimal breeding habitat for these mosquitoes, facilitating the replication of dengue viruses.

**Fig 1.**
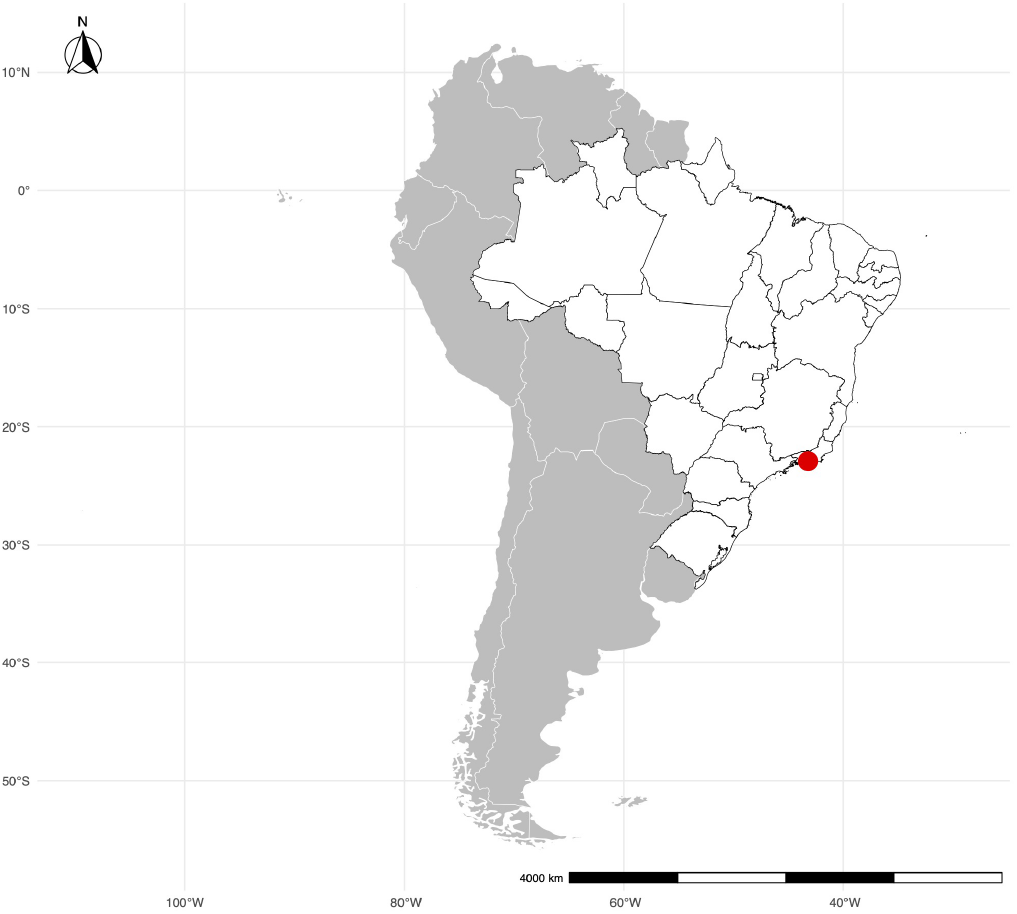
Map of South America with Rio de Janeiro marked by a red point.

Dengue is a notifiable disease within Brazil’s public health framework, requiring health workers to report suspected cases through a structured notification process. This process culminates in a national database managed by the Ministry of Health, which, despite the limitation that only a fraction of cases are laboratory-confirmed, provides invaluable incidence indicators for disease monitoring and response.

The InfoDengue system [22] offers a semi-automated pipeline for data collection, harmonization, and analysis at the municipal level. This system generates crucial indicators of the epidemiological situation of dengue and other arboviruses such as Zika and Chikungunya, facilitating timely and informed public health responses. In addition to the reported disease cases, InfoDengue incorporates weather data, acknowledging the significant impact of climate on arbovirus transmission. Specifically, temperature and humidity data, sourced from airport weather stations and satellite imagery, are provided to understand and predict disease spread patterns.

Figure 2 displays the weekly cases of dengue reported in Rio de Janeiro from 2016 to 2023. As seen in the graph, dengue cases display significant variability with an average of approximately 295 cases, and notable peaks reaching up to 3127 cases. The months with higher dengue cases typically occur between March and May, consistent with known seasonal patterns in Brazil [37–39]. Figure 3 illustrates the patterns of median weekly temperature and humidity over the same period. Notably, temperature and humidity exhibit opposite trends, when temperature is high, humidity tends to be low, and vice versa. The median weekly temperature over the study period was 23.15°C, with values ranging from 15.60°C to 30.55°C. Humidity levels showed an average of 79.36%, fluctuating between 54.20% and 91.90%, reflecting seasonal variations. Temperature tends to peak during the summer months, while humidity shows some variability across seasons. The fluctuations in these climate factors often align with changes in dengue cases, underscoring their importance in predicting disease transmission.

**Fig 2.**
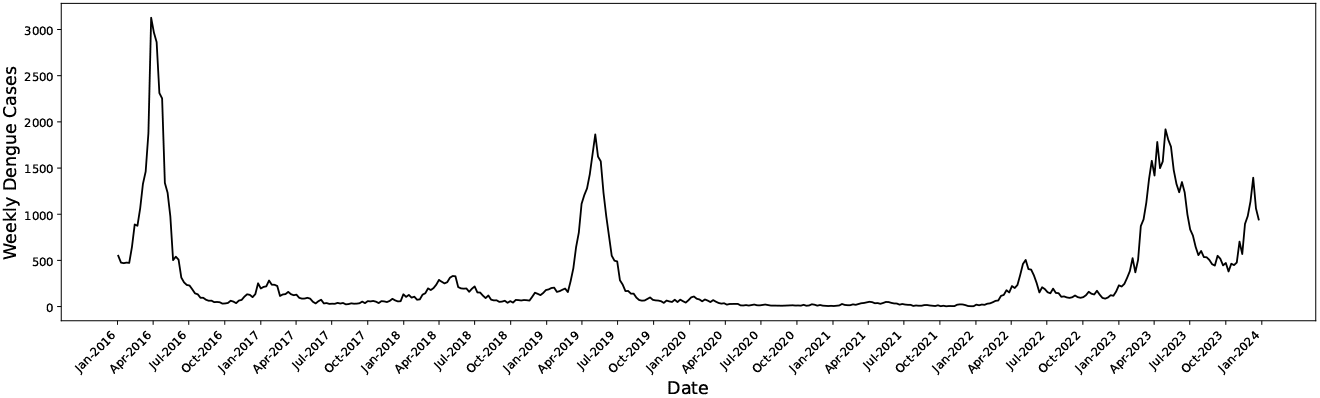
Dengue cases per week in Rio de Janeiro, Brazil.

**Fig 3.**
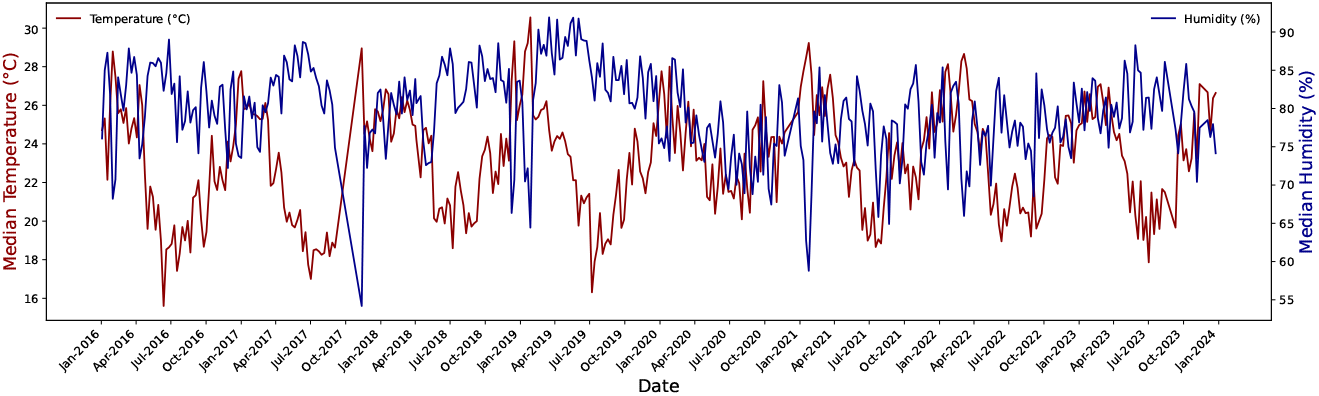
Weekly temperature and humidity in Rio de Janeiro, Brazil.

## Methodology

We compare the predictive performance and computational time of a number of statistical methods and machine learning techniques to forecast dengue cases using a moving window strategy. In this strategy, also known as a rolling forecast or rolling window approach, the model is trained on a fixed-size segment of historical data to predict future values. As new data becomes available, the window moves forward by one or more time points, dropping the oldest data in the set and incorporating the most recent data for subsequent predictions. This method is particularly advantageous for forecasting tasks where the relationship between past and future values may change over time.

For the purpose of predicting dengue cases in Rio de Janeiro, we implemented a moving window strategy with a fixed window size of 6 years (Figure 4). This window size was selected to capture the long-term trends and seasonal patterns of dengue cases while providing a sufficiently large dataset for model training. Initially, the training window consisted of 2016-01-03 to 2021-12-26 and was moved one week until 2023-12-24. The forecasting horizon was set to 1, 2, 3, 4, 8, and 12 weeks ahead, allowing us to evaluate the models’ performance over short to medium-term predictions. In the following sections, we describe the statistical methods and machine learning techniques employed, the procedure to compute the associated uncertainty intervals, and the performance evaluation metrics utilized for comparison.

**Fig 4.**
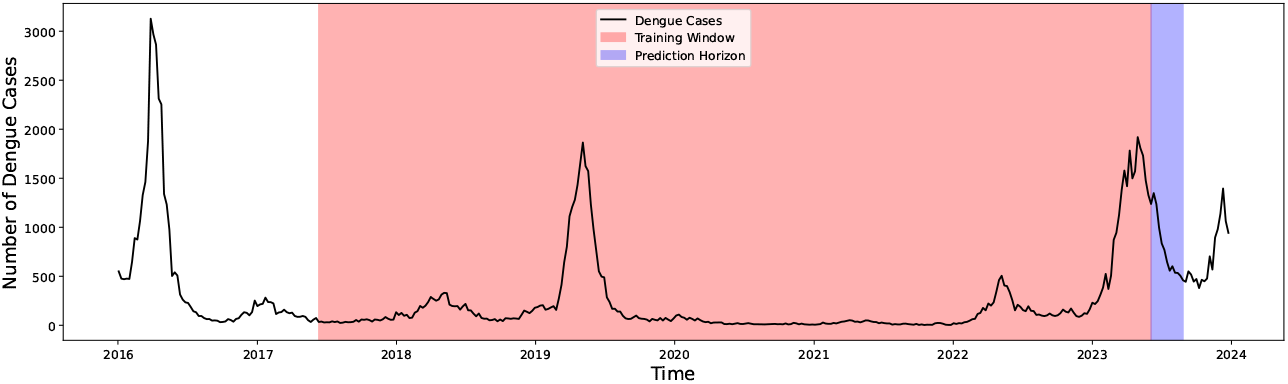
Illustration of the moving window strategy for dengue case forecasting.

### Statistical models for dengue forecasting

The statistical models considered to predict dengue include Autoregressive (AR(1)), Moving Average (MA(1)), Autoregressive Integrated Moving Average (ARIMA) [23, 24], and Exponential Smoothing State Space Model (ETS) [25]. These models were chosen for their ability to capture various patterns in time series data, such as trends, seasonality, and autocorrelation. In addition, we use models that incorporate temperature and humidity as covariates to capture the influence of these factors on dengue transmission. These models are Vector Autoregression (VAR) [23, 26] and Seasonal ARIMAX (SARIMAX) [24]. Furthermore, to account for the delayed effects of climatic factors on dengue incidence, VAR and SARIMAX models are also fitted using lagged variables ranging from 1 week to 4 weeks. Previous studies support this approach, indicating that temperature and humidity at a lag of one month are positively associated with dengue incidence [40, 41]. Let *X*_*t*_ represent the number of dengue cases at time *t*. The statistical models employed are specified as follows:

- The Autoregressive Model (AR(1)) predicts future values based on a combination of past values. It is defined as

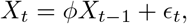

where *ϕ* is the coefficient that measures the influence of the immediately preceding value (*X*_*t*−1_), and *ϵ*_*t*_ is the white noise error term at time *t*. This model is particularly effective for data showing a strong correlation with its immediate past value.
- The Moving Average Model (MA(1)) model captures the relationship between an observation and a residual error from a moving average model applied to lagged observations. It is represented as

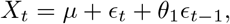

where *µ* is the mean of the series, *ϵ*_*t*_ is the white noise error term, and *θ*_1_ is the coefficient for the lagged error term. This model is effective in smoothing out short-term fluctuations and identifying patterns that persist over time.
- The Autoregressive Integrated Moving Average (ARIMA) model combines the AR and MA models and integrates differencing to make the data stationary. It is denoted as ARIMA(*p, d, q*), where *p* is the order of the AR term, *d* is the degree of differencing, and *q* is the order of the MA term. The general form of an ARIMA model is

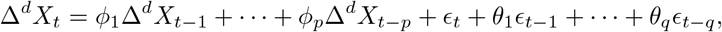

where Δ^*d*^ denotes differencing *d* times to achieve stationarity. ARIMA models are versatile and can model data with trends and seasonal components.
- The Exponential Smoothing State Space Model (ETS) model accounts for trends and seasonality in the data through exponential smoothing. The general form of the ETS model can be written as

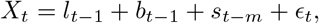

where *l*_*t*_ is the level of the series, capturing the long-term average behavior; *b*_*t*_ is the trend, indicating the direction and speed of the change; *s*_*t*_ is the seasonal component, capturing periodic fluctuations; *m* is the period of seasonality, and *ϵ*_*t*_ is the error term. ETS models are particularly useful for capturing complex seasonality patterns in time series data.
- The VAR model captures the linear interdependencies among multiple time series. It generalizes the AR model by allowing for more than one evolving variable. All the variables in a VAR are treated symmetrically; each variable has an equation explaining its evolution based on its own lags and the lags of the other model variables. For a system of *n* time series (in our case, dengue cases, temperature, and humidity), a VAR model of order *p* (VAR(*p*)) can be written as follows:

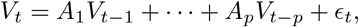

where *V*_*t*_ is a vector of time series variables including dengue cases, temperature, and humidity at time *t, A*_*i*_ are coefficient matrices, and *ϵ*_*t*_ is a vector of error terms. VAR models are suitable for capturing the dynamic relationships between multiple time series, such as dengue cases, temperature, and humidity.
- Seasonal ARIMAX (SARIMAX) extends the ARIMA model by incorporating exogenous variables (X) and seasonal components (S). It is a powerful and flexible model that can account for complex behaviors in time series data. It is represented as SARIMAX(p, d, q)(P, D, Q) s, where *p, d, q* are the non-seasonal parameters, *P, D, Q* are the seasonal parameters, and *s* is the length of the seasonal cycle. Let *X*_*t*_ be the number of dengue cases at time *t*, and let *Z*_*t*_ be the vector of exogenous variables (i.e., temperature and humidity), the SARIMAX model can be represented as

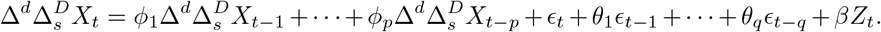

SARIMAX is particularly effective in incorporating seasonal effects and external influences on dengue transmission.

### Machine learning techniques for dengue forecasting

We also consider various machine learning techniques that have demonstrated significant potential in enhancing dengue forecasting by utilizing both historical dengue case data and additional climate covariates such as temperature and humidity. Our selection includes a diverse array of models, each chosen for their unique strengths in dealing with complex epidemiological data, and their flexibility allows for the integration of lagged variables, providing a richer analysis that accounts for past data trends and environmental influences. These methods include decision tree-based models like Random Forest and XGBoost, as well as Support Vector Machine (SVM), Long Short-Term Memory (LSTM) networks, and Prophet.

For this study, we utilize these machine learning methods in two scenarios: using only historical dengue case data, and incorporating temperature and humidity as covariates. Similar to the statistical methods, we also incorporate lagged variables from 1 to 4 weeks to capture delayed effects of climatic factors on dengue incidence. The machine learning methods are specified as follows:

- Random Forest [27], known for its simplicity and effectiveness, it is an ensemble learning method that constructs multiple decision trees during training and outputs the average prediction of the individual trees. Each tree in the forest is built from a bootstrap sample of the training data, and at each split, a random subset of features is considered. This method is effective in reducing overfitting and improving generalization. For dengue forecasting, Random Forest can model historical case data, making it suitable for capturing the stochastic nature of dengue transmission.
- Extreme Gradient Boosting (XGBoost) [28] is an advanced implementation of gradient-boosted decision trees, which has gained popularity due to its speed and performance, which stems from its ability to do gradient boosting in a more efficient way, making it particularly suited for large datasets. It builds trees sequentially, where each subsequent tree focuses on correcting the errors of the previous trees. This method enhances model accuracy and robustness by combining multiple weak learners into a strong learner. XGBoost’s ability to handle missing values and incorporate regularization makes it well-suited for dengue forecasting.
- Support Vector Machine (SVM) [29, 30] for regression, known as Support Vector Regression (SVR), attempts to find a function that deviates from the actual observed values by a value no greater than a specified margin while balancing the complexity of the model. SVR is particularly effective for capturing non-linear relationships in the data. When applied to dengue forecasting, SVR can model the complex and non-linear interactions between historical dengue cases and climatic factors.
- Long Short-Term Memory (LSTM) networks [31, 32] is a type of recurrent neural network capable of learning order dependence in sequence prediction problems. LSTM cells have internal mechanisms called gates that regulate the flow of information, making them highly effective for learning from sequences of data, such as weekly dengue case reports, where past information is crucial for predicting future events. LSTM networks can learn the temporal patterns in dengue case data and the influence of lagged climatic factors such as temperature and humidity, which allows LSTMs to forecast future dengue incidences with enhanced accuracy.
- Prophet [33] is a forecasting tool that models time series data with strong seasonal effects. It is a procedure for forecasting time series data based on an additive model, decomposing the time series into trend, seasonality, and holiday effects. It works well with time series that have strong seasonal effects and historical trends, making it ideal for predicting dengue cases because of its robust handling of seasonal variations and its capability to model non-linear trends influenced by yearly and weekly cycles of dengue cases. By adjusting its parameters, Prophet can also integrate additional regressors such as temperature and humidity, aligning closely with the seasonal patterns that affect dengue transmission.

### Ensemble approaches

Aiming to enhance the accuracy and stability of dengue prediction techniques, we also employed ensemble approaches that averaged the forecast outputs from both the best-performing statistical models and machine learning techniques independently trained on historical data. This approach is designed to harness the complementary strengths of each model type, namely, the statistical model’s efficacy in capturing linear trends and seasonality, and the machine learning model’s ability to understand complex, non-linear relationships in data sequences. Thus, the ensemble approaches mitigate individual model prediction errors, potentially reducing the overall forecast variance without excessively complicating the model structure.

### Uncertainty intervals

Adaptive conformal prediction [42–45] is a technique used to construct prediction intervals that account for time-dependent changes in the data distribution, making it particularly suitable for time series data. This method adjusts the prediction intervals dynamically based on nonconformity scores to provide more accurate and reliable intervals. We use adaptive conformal prediction to compute uncertainty intervals associated to each of the methods under consideration.

For each time *t* and forecast horizon *h*, the nonconformity score is defined as the residual at time *t* for *h* steps ahead calculated as the difference between the actual and predicted values from the time series model:

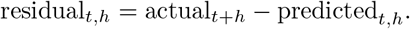

For a window size *W* and time *t*, residuals are considered from *t* − *W* to *t*. Thus, the set of nonconformity scores within the window at time *t* for a forecast horizon *h* is

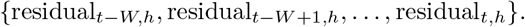

The quantiles of these nonconformity scores are then computed to determine the prediction intervals. Specifically, the lower and upper bounds of the prediction interval at each time step *t* for forecast horizon *h* are given by

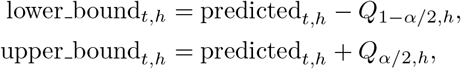

where *Q*_1−*α/*2,*h*_ and *Q*_*α/*2,*h*_ are the quantiles of the residuals within the window for the *h*-step ahead forecast, and *α* is the significance level (e.g., 0.05 for a 95% prediction interval). For the *k*-th quantile at forecast horizon *h*, the corresponding value is given by

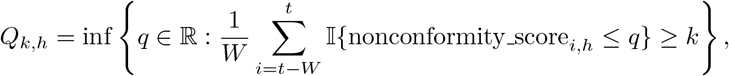

where 𝕀 is the indicator function.

The prediction interval is then adjusted dynamically at each time step based on these scores. This adaptive update ensures that the prediction intervals remain accurate as the underlying data distribution evolves, providing more reliable intervals that can better account for time-dependent changes in the data.

### Methods implementation

Statistical models AR(1), MA(1), ARIMA, SARIMAX, and ETS are implemented using the `R` package `forecast` [46], while the VAR model is implemented using the `R` package `vars` [47]. Machine learning models Random Forest and XGBoost are implemented using the `Python` libraries `scikit-learn` [48] and `xgboost` [28], respectively. SVM is implemented using `scikit-learn`, LSTM networks are implemented using the `TensorFlow` library [49], and Prophet is implemented using the `prophet` library [50]. For reproducibility purposes, we provide a GitHub repository with the codes, which can be found at https://github.com/ChenXiang1998/Assessing-Dengue-Forecasting-Methods.

### Performance evaluation metrics

To assess the predictive performance of the statistical models and machine learning techniques considered, we employed four primary metrics, namely, Mean Absolute Error (MAE), Mean Absolute Percentage Error (MAPE), Root Mean Squared Error (RMSE), and 95% Coverage Probability. These metrics provide a comprehensive view of the performance of the models, considering both the magnitude and direction of prediction errors, and help highlight the strengths and weaknesses in dengue forecasting methods. In addition, we also compare the computational efficiency of the forecasting techniques considered by measuring the duration that a technique takes to complete its training and prediction processes. This comparison provides insights on the practicality of deploying each method in real-world situations, where computational resources and response times are often limited. It also assists public health workers on which models to deploy, balancing the need between predictive accuracy and computational efficiency.

Let *y*_*i*_ and *ŷ*_*i*_ represent, respectively, the actual and forecast number of dengue cases, and let *n* be the number of observations. The Mean Absolute Error (MAE) measures the average magnitude of the errors in a set of predictions, without considering their direction. It is calculated as the average of the absolute differences between the forecasted values and the actual values:

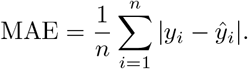

The Mean Absolute Percentage Error (MAPE) expresses the accuracy as a percentage, which provides a relative measure of the errors. It is calculated as the average of the absolute percentage differences between the predicted and actual values:

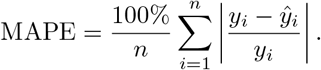

The Root Mean Squared Error (RMSE) is a quadratic scoring rule that measures the average magnitude of the error. It is defined as the square root of the average of squared differences between prediction and actual observation:

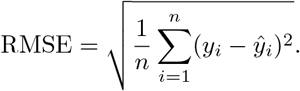

The 95% Coverage Probability assesses the proportion of actual dengue cases that fall within the 95% prediction intervals provided by the models. A well-calibrated model should have approximately 95% of the observations within this interval. It is a measure of the reliability of the predictive intervals.

## Results

In this section, we evaluate the predictive performance of the statistical models, machine learning techniques, and ensemble approaches considered. We provide the methods’ predictive accuracy in terms of MAE, RMSE, and MAPE across various forecast horizons. Then, we provide the 95% coverage probability and the average width of the uncertainty intervals of all models across various forecast horizons. A comparison of the computational efficiency of each method is also provided. In the Supporting information, Figure S1 shows a comparison of the predictions obtained by the best performing methods for different time horizons. Figures S2 to S5 depict boxplots of the absolute error, |*y*_*i*_ − *ŷ*_*i*_|, and absolute percentage errors, 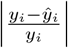, obtained with each method. Specifically, Figure S2 and Figure S3 correspond to methods that only use dengue cases, while Figure S4 and Figure S5 show boxplots for the methods using covariates. In all figures, boxplots are shown in the order from best to worst approach forecasting method.

### Performance of statistical models

Table 1 shows the accuracy metrics for the statistical models that did not include covariates, namely, AR(1), MA(1), ARIMA, and ETS. Among these models, ARIMA stands out as the most consistent performer across varying forecast horizons. Its ability to blend autoregressive (AR), differencing (I), and moving average (MA) components allows it to capture both short-term trends and long-term seasonal patterns effectively.

**Table 1.** Accuracy of the statistical models without covariates at various forecast horizons.

| Week | AR(1) |  |  | MA(1) |  |  | ARIMA |  |  | ETS |  |  |
| --- | --- | --- | --- | --- | --- | --- | --- | --- | --- | --- | --- | --- |
|  | MAE | MAPE | RMSE | MAE | MAPE | RMSE | MAE | MAPE | RMSE | MAE | MAPE | RMSE |
| 1 | 83.28 | 19.93% | 127.03 | 82.72 | 19.49% | 124.61 | **78.26** | **18.25%** | **124.33** | 78.66 | 19.17% | 126.71 |
| 2 | 114.99 | 26.59% | 176.96 | 114.62 | 26.03% | **175.04** | **114.61** | **25.05%** | 179.33 | 118.36 | 26.52% | 185.13 |
| 3 | **139.51** | 33.02% | **211.01** | 144.65 | 32.90% | 216.32 | 143.84 | **31.39%** | 221.81 | 146.24 | 36.07% | 223.62 |
| 4 | 178.87 | 39.48% | **269.14** | 182.26 | 39.85% | 274.49 | **175.38** | **36.07%** | 277.48 | 186.66 | 44.12% | 292.18 |
| 8 | 314.44 | 62.00% | **457.62** | 323.80 | 62.68% | 465.38 | **308.92** | **56.32%** | 461.73 | 392.21 | 89.16% | 578.60 |
| 12 | 432.34 | 81.05% | 608.53 | 436.55 | 81.86% | 612.83 | **396.91** | **64.95%** | **583.04** | 649.15 | 150.31% | 917.47 |

For 1-week ahead forecasts, ARIMA exhibited the lowest MAE at 78.26 cases and the lowest MAPE at 18.25%, indicating its superior performance for immediate forecasting compared to other models. The ETS model closely followed, with an MAE of 78.66 cases and a slightly higher MAPE of 19.17%. The AR(1) and MA(1) models showed slightly higher errors but were competitive in their forecasting ability. In terms of RMSE, ARIMA also had the lowest value (124.33), making it the most accurate at this horizon.

Extending the forecast horizon to 4 weeks ahead, we observed an increase in the errors across all models, as expected due to the increasing uncertainty with longer forecast periods. ARIMA maintained a relatively lower MAE and MAPE, affirming its consistency in performance across the short-term forecast horizons. For instance, ARIMA’s MAE and MAPE at 4 weeks were 175.38 and 36.07%, respectively, which were the best among the considered models.

As the forecast horizon expanded to 8 and 12 weeks, all models’ performance naturally deteriorated, reflecting the inherent challenge of predicting dengue cases over longer periods. Notably, the ARIMA model’s performance remained robust relative to the other models, with the lowest MAE of 308.92 and 396.91 for 8 and 12 weeks ahead, respectively. However, the ETS model’s error metrics significantly increased, with a notable jump to 392.21 (MAE) and 89.16% (MAPE) at 8 weeks and 649.15 (MAE) and 150.31% (MAPE) at 12 weeks, suggesting that it may be less suitable for medium- and long-term forecasting in the absence of covariates.

In an effort to enhance the predictive accuracy of the statistical models, we considered SARIMAX and VAR models that included temperature and humidity covariates, both with and without lagged covariates. Results corresponding to these models are shown in Table 2.

**Table 2.** Accuracy of the statistical models with covariates temperature and humidity at various forecast horizons.

| week | SARIMAX |  |  | SARIMAX_Lag |  |  | VAR |  |  | VAR_Lag |  |  |
| --- | --- | --- | --- | --- | --- | --- | --- | --- | --- | --- | --- | --- |
|  | MAE | MAPE | RMSE | MAE | MAPE | RMSE | MAE | MAPE | RMSE | MAE | MAPE | RMSE |
| 1 | 79.24 | **17.25%** | 128.36 | 81.80 | 25.23% | 125.36 | 78.10 | 25.49% | 123.49 | **77.74** | 24.44% | **116.24** |
| 2 | 114.63 | **24.27%** | 181.58 | 114.74 | 36.96% | 167.61 | 118.08 | 40.70% | 175.92 | **108.63** | 35.11% | **166.80** |
| 3 | 143.39 | **30.85%** | 222.82 | **136.62** | 39.64% | **200.87** | 148.71 | 48.08% | 214.39 | 137.41 | 43.72% | 207.91 |
| 4 | 173.52 | 36.59% | 278.54 | **160.28** | 39.96% | **246.61** | 179.45 | 55.57% | 267.68 | 167.53 | 48.79% | 259.62 |
| 8 | 316.86 | 57.84% | 473.41 | **279.15** | **54.86%** | **414.65** | 313.80 | 65.08% | 444.61 | 295.42 | 55.03% | 438.32 |
| 12 | 408.08 | 67.88% | 598.16 | **375.15** | 73.33% | **536.71** | 391.64 | 64.77% | 566.66 | 386.34 | **62.41%** | 579.80 |

In the short-term forecast horizon of 1 week, SARIMAX outperformed others, achieving the lowest MAPE of 17.25% and an MAE of 79.24 cases. When considering lagged covariates, SARIMAX Lag’s performance slightly declined, with an MAE of 81.80 cases and a MAPE of 25.23%. VAR showed competitive performance with an MAE of 78.10 cases but a higher MAPE of 25.49%.

As the horizon extends to 4 weeks, the predictive accuracy decreased across all models. SARIMAX continued to demonstrate strong performance, with an MAE of 173.52 cases and a MAPE of 36.59%. The inclusion of lagged covariates in SARIMAX Lag resulted in an MAE of 160.28 and a MAPE of 39.96%, indicating a slight performance degradation. VAR exhibited an MAE of 179.45 cases and a MAPE of 55.57%, while VAR Lag had an MAE of 167.53 and a MAPE of 48.79%.

For 8 and 12 weeks ahead forecasts, the performance of all models continued to decline. SARIMAX Lag, however, consistently maintained a lower error rate relative to other models, with an MAE of 279.15 and 375.15 and an MAPE of 54.86% and 73.33% for 8 and 12 weeks ahead, respectively. VAR showed increased errors, with an MAE of 313.80 and 386.34 for 8 and 12 weeks, respectively. The VAR Lag model’s performance was comparatively better, with an MAE of 295.42 and 386.34 for the same horizons.

Overall, incorporating temperature and humidity covariates improved the models’ predictive accuracy. The SARIMAX model, in particular, demonstrated superior performance compared to ARIMA, indicating that these covariates are valuable for short-term predictions. Additionally, models incorporating lagged variables, such as SARIMAX Lag and VAR Lag, showed enhanced predictive capability for medium- and long-term forecasts. This suggests that lagged variables can significantly improve the accuracy of dengue case predictions over longer periods.

### Performance of machine learning techniques

In this section, we describe the performance of the five machine learning techniques considered, namely, SVM, Random Forest, XGBoost, LSTM and Prophet. Table 3 shows the accuracy measures MAE, RMSE, and MAPE across various forecast horizons for the machine learning techniques that did not use covariates. Table 4 shows the accuracy metrics for the machine learning techniques that included temperature and humidity covariates. The results are provided for tecniques that used covariates for the same week as well as for lagged covariates.

**Table 3.**
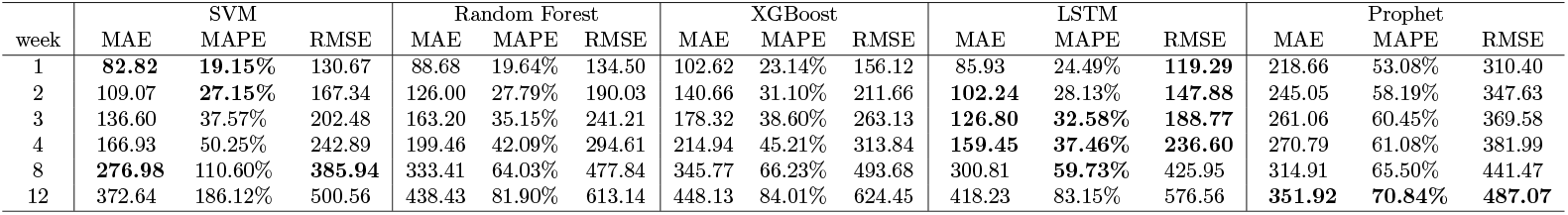
Accuracy of the machine learning techniques considered without covariates at various forecast horizons.

**Table 4.** Accuracy of the machine learning techniques considered with covariates temperature and humidity at various forecast horizons.

| week | SVM |  |  | Random Forest |  |  | XGBoost |  |  | LSTM |  |  | Prophet |  |  |
| --- | --- | --- | --- | --- | --- | --- | --- | --- | --- | --- | --- | --- | --- | --- | --- |
|  | MAE | MAPE | RMSE | MAE | MAPE | RMSE | MAE | MAPE | RMSE | MAE | MAPE | RMSE | MAE | MAPE | RMSE |
| 1 | 112.29 | 29.84% | 176.15 | 410.44 | 119.94% | 621.41 | 439.89 | 151.00% | 644.90 | **71.35** | **22.31%** | **101.53** | 200.20 | 46.62% | 293.64 |
| 2 | 138.34 | 36.41% | 215.68 | 418.29 | 119.06% | 628.82 | 440.81 | 139.93% | 646.74 | **89.50** | **23.56%** | **130.71** | 222.88 | 49.96% | 326.45 |
| 3 | 165.67 | 46.73% | 248.43 | 416.61 | 80.38% | 628.41 | 431.51 | 93.65% | 636.03 | **114.73** | **26.24%** | **173.90** | 247.76 | 54.57% | 353.17 |
| 4 | 193.32 | 57.77% | 280.87 | 425.16 | 79.53% | 635.42 | 452.01 | 92.57% | 653.74 | **148.74** | **32.14%** | **223.35** | 265.58 | 57.98% | 373.31 |
| 8 | **289.62** | 108.35% | **405.34** | 444.25 | 69.99% | 652.34 | 461.87 | 78.97% | 660.34 | 292.91 | **57.01%** | 416.62 | 316.96 | 64.81% | 444.14 |
| 12 | 360.53 | 175.58% | **492.24** | 463.77 | 69.96% | 667.65 | 465.48 | 73.89% | 665.71 | 410.96 | 79.92% | 573.32 | **342.47** | **61.86%** | 495.31 |

| week | SVM-Lag |  |  | Random Forest-Lag |  |  | XGBoost-Lag |  |  | LSTM-Lag |  |  | Prophet-Lag |  |  |
| --- | --- | --- | --- | --- | --- | --- | --- | --- | --- | --- | --- | --- | --- | --- | --- |
|  | MAE | MAPE | RMSE | MAE | MAPE | RMSE | MAE | MAPE | RMSE | MAE | MAPE | RMSE | MAE | MAPE | RMSE |
| 1 | 188.99 | 50.10% | 310.12 | 414.14 | 143.38% | 603.79 | 412.62 | 172.51% | 610.86 | 83.58 | 28.87% | 117.39 | 208.55 | 77.77% | 299.80 |
| 2 | 205.94 | 56.69% | 338.36 | 422.68 | 142.14% | 613.06 | 433.29 | 210.35% | 627.08 | 102.1 | 28.27% | 155.82 | 235.76 | 65.11% | 339.18 |
| 3 | 223.76 | 63.06% | 364.19 | 424.42 | 117.13% | 616.01 | 416.41 | 145.61% | 611.33 | 132.8 | 30.88% | 201.65 | 259.27 | 58.71% | 369.47 |
| 4 | 245.03 | 70.38% | 391.97 | 428.85 | 109.80% | 621.38 | 427.03 | 154.94% | 612.99 | 166.45 | 34.97% | 251.28 | 280.24 | 59.53% | 394.74 |
| 8 | 303.25 | 85.87% | 470.16 | 439.87 | 71.77% | 641.60 | 431.1 | 74.68% | 630.37 | 304.93 | 60.16% | 438.11 | 347.77 | 70.88% | 479.95 |
| 12 | **341.82** | 121.50% | 513.45 | 454.75 | 68.21% | 655.17 | 446.64 | 71.04% | 646.80 | 422.09 | 82.63% | 586.77 | 389.37 | 74.56% | 540.32 |

For 1-week ahead forecasts, SVM exhibited the lowest MAE at 82.82 cases and the lowest MAPE at 19.15%, indicating its strong short-term predictive ability. LSTM also performed well, with an MAE of 85.93 cases and a MAPE of 24.49%, showing its competitive edge for immediate forecasting. Random Forest and XGBoost showed slightly higher errors but were still effective, with MAEs of 88.68 and 102.24 cases, respectively. Prophet, however, showed much higher errors, with an MAE of 218.66 cases and a MAPE of 53.08%, indicating its limitations in short-term forecasting.

Extending the forecast horizon to 4 weeks, we observed an increase in the errors across all models. LSTM model demonstrated the lowest MAE at 159.45 cases and a MAPE of 37.46%, indicating its robustness in medium-term forecasting. SVM, while having a higher MAE of 166.93, still maintained a relatively lower MAPE of 50.25%. XGBoost showed an MAE of 199.46 cases and a MAPE of 42.09%, performing better than Random Forest and Prophet at this horizon. Prophet continued to exhibit higher errors with an MAE of 270.79 cases and a MAPE of 61.08%, reinforcing its unsuitability for medium-term forecasts.

For 8 and 12 weeks ahead forecasts, all models’ performance naturally deteriorated due to the increasing uncertainty over longer periods. LSTM continued to show the lowest errors with an MAE of 300.81 for 8 weeks ahead. However, for the 12-week horizon, Prophet exhibited the best performance with the lowest MAE of 351.92 cases and a MAPE of 70.84%. This indicates that while Prophet is not suitable for short-term forecasts, it shows strong predictive capabilities for very long-term forecasts.

Overall, the machine learning models’ performance varied across different forecast horizons, with SVM and LSTM models showing superior accuracy for short-term and medium-term predictions, respectively. Prophet, despite its poor short-term performance, demonstrated notable effectiveness in long-term forecasting.

Incorporating covariates, LSTM was the best performer, achieving the lowest MAE of 71.35 cases and a MAPE of 22.31% for 1-week ahead forecasts. This model continued to show the best performance across all forecast horizons up to 12 weeks, with an MAE of 410.96 cases and a MAPE of 79.92%. However, the other models showed significantly higher errors when covariates were included, particularly the tree-based models such as Random Forest and XGBoost. For example, Random Forest with covariates had an MAE of 410.44 and a MAPE of 119.94% for 1-week ahead, which did not improve substantially over longer horizons.

Lagged covariates negatively impacted the performance of all machine learning models across all forecast horizons, worsening MAE, MAPE and RMSE values, indicating potential overfitting due to the redundant of lag data. The lagged models, including SVM-Lag, Random Forest-Lag, XGBoost-Lag, LSTM-Lag, and Prophet-Lag, exhibited higher errors across all forecast horizons compared to their counterparts without lagged covariates. For instance, SVM-Lag had an MAE of 188.99 cases and a MAPE of 50.10% for 1-week ahead, with the performance deteriorating over longer horizons to an MAE of 341.82 and a MAPE of 121.50% at 12 weeks. Similarly, the LSTM-Lag model, despite having some competitive metrics for short-term forecasts, showed significant error increases for longer horizons.

Prophet with covariates showed improved performance compared to its counterpart without covariates, particularly for long-term forecasts. For 12 weeks ahead, Prophet with covariates had an MAE of 342.47 and a MAPE of 61.86%, making it effective for very long-term predictions. However, like other models, its short-term performance remained relatively weak.

Overall, while incorporating temperature and humidity covariates improved the LSTM model’s predictive accuracy significantly, other models did not benefit similarly and, in many cases, performed worse. Additionally, the use of lagged covariates generally led to poorer performance, suggesting that these models may suffer from overfitting when lagged variables are included. Prophet with covariates, although not performing well in the short-term, continued to show strong predictive capabilities for very long-term forecasts.

In summary, LSTM including covariates dominated the short-to mid-term horizons (i.e., 1 to 8 weeks), providing consistently low errors across these intervals. Prophet proved to be a reliable model for long-term forecasting, excelling at 12 weeks with its ability to identify and capture longer-term patterns in dengue case data. LSTM and Prophet demonstrate complementary strengths. Specifically, LSTM is highly effective for immediate and mid-term predictions, while Prophet is better suited for capturing long-term trends.

### Performance ensemble approaches

In this section, we discuss the performance of the ensemble models, which combine statistical methods and machine learning approaches to enhance predictive accuracy. The choice of models for the ensembles was based on their individual performance metrics. By averaging the best-performing statistical models and machine learning models, we aimed to leverage the strengths of each model.

For predictions using dengue cases alone, we created an ensemble model by averaging the forecasts of the ARIMA and LSTM models. Table 5 presents the performance metrics for this ensemble model compared to the individual ARIMA and LSTM models.

**Table 5.** Accuracy of ensemble model of LSTM & ARIMA compared with ARIMA and LSTM without covariates at various forecast horizons.

| week | LSTM-ARIMA |  |  | ARIMA |  |  | LSTM |  |  |
| --- | --- | --- | --- | --- | --- | --- | --- | --- | --- |
|  | MAE | MAPE | RMSE | MAE | MAPE | RMSE | MAE | MAPE | RMSE |
| 1 | **69.98** | **17.38%** | **108.27** | 78.26 | 18.25% | 124.33 | 85.93 | 24.49% | 156.12 |
| 2 | **97.10** | **21.89%** | **148.67** | 114.61 | 25.05% | 179.33 | 102.24 | 28.13% | 211.66 |
| 3 | **126.15** | **27.54%** | **189.53** | 143.84 | 31.39% | 221.81 | 126.80 | 32.58% | 263.13 |
| 4 | **154.43** | **32.19%** | **242.39** | 175.38 | 36.07% | 277.48 | 159.45 | 37.46% | 313.84 |
| 8 | **286.53** | **52.97%** | **427.47** | 308.92 | 56.32% | 461.73 | 300.81 | 59.73% | 493.68 |
| 12 | **379.67** | 67.79% | **555.60** | 396.91 | **64.95%** | 583.04 | 418.23 | 83.15% | 624.45 |

The LSTM-ARIMA ensemble shows significant improvements over the individual ARIMA and LSTM models. For instance, for 1-week ahead forecasts, the ensemble model achieved the lowest MAE of 69.98 cases and a MAPE of 17.38%. This outperformed both ARIMA and LSTM models individually, which had MAEs of 78.26 and 85.93 cases, respectively. The ensemble model also exhibited the lowest RMSE of 108.27, indicating enhanced predictive accuracy.

As the forecast horizon extends, the LSTM-ARIMA ensemble continues to demonstrate superior performance, particularly noticeable at 4 weeks ahead, where the ensemble’s MAE and MAPE were 154.43 cases and 32.19%, respectively, compared to higher errors in the individual models. Although the performance of all models declines over longer horizons, the ensemble approach still maintains a comparative advantage, highlighting its robustness in dengue case forecasting.

For predictions including temperature and humidity covariates, we created several ensemble models. These included combinations of SARIMAX, SARIMAX-Lag, and VAR-Lag with LSTM, as well as a more complex ensembles LSTM-SARIMAX-VAR Lag and LSTM-SARIMAX-Lag-VAR-Lag. Table 6 presents the performance metrics for these ensemble models compared to the individual models.

**Table 6.** Accuracy of ensemble model of LSTM, SARIMAX & VAR compared with the individual models with covariates temperature and humidity at various forecast horizons.

| week | SARIMAX |  |  | SARIMAX-Lag |  |  | VAR-Lag |  |  | LSTM |  |  |
| --- | --- | --- | --- | --- | --- | --- | --- | --- | --- | --- | --- | --- |
|  | MAE | MAPE | RMSE | MAE | MAPE | RMSE | MAE | MAPE | RMSE | MAE | MAPE | RMSE |
| 1 | 79.24 | 17.25% | 128.36 | 81.80 | 25.23% | 125.36 | 77.74 | 24.44% | 116.24 | 71.35 | 22.31% | 101.53 |
| 2 | 114.63 | 24.27% | 181.58 | 114.74 | 36.96% | 167.61 | 108.63 | 35.11% | 166.8 | 89.50 | 23.56% | **130.71** |
| 3 | 143.39 | 30.85% | 222.82 | 136.62 | 39.64% | 200.87 | 137.41 | 43.72% | 207.91 | 114.73 | 26.24% | **173.90** |
| 4 | 173.52 | 36.59% | 278.54 | 160.28 | 39.96% | 246.61 | 167.53 | 48.79% | 259.62 | 148.74 | 32.14% | **223.35** |
| 8 | 316.86 | 57.84% | 473.41 | 279.15 | 54.86% | 414.65 | 295.42 | 55.03% | 438.32 | 292.91 | 57.01% | 416.62 |
| 12 | 408.08 | 67.88% | 598.16 | **375.15** | 73.33% | **536.71** | 386.34 | **62.41%** | 579.8 | 410.96 | 79.92% | 573.32 |

| week | LSTM-SARIMAX |  |  | LSTM-SARIMAX-Lag |  |  | LSTM-VAR-Lag |  |  | LSTM-SARIMAX-VAR-Lag |  |  | LSTM-SARIMAX-Lag-VAR-Lag |  |  |
| --- | --- | --- | --- | --- | --- | --- | --- | --- | --- | --- | --- | --- | --- | --- | --- |
|  | MAE | MAPE | RMSE | MAE | MAPE | RMSE | MAE | MAPE | RMSE | MAE | MAPE | RMSE | MAE | MAPE | RMSE |
| 1 | 65.24 | **15.82%** | 103.52 | 66.17 | 18.83% | 102.59 | 57.92 | 20.83% | **89.73** | **57.86** | 17.36% | 94.03 | 59.71 | 20.19% | 95.23 |
| 2 | 92.17 | **18.69%** | 144.27 | 93.98 | 25.89% | 141.95 | **85.68** | 26.23% | 133.39 | 87.84 | 23.09% | 139.12 | 90.22 | 28.87% | 138.29 |
| 3 | 118.90 | **24.11%** | 184.84 | 118.90 | 28.64% | 180.90 | 113.80 | 29.96% | 178.01 | **113.00** | 25.87% | 183.03 | 114.20 | 31.37% | 180.19 |
| 4 | 147.79 | **29.89%** | 238.06 | 148.80 | 31.88% | 229.42 | 146.64 | 34.63% | 228.74 | **141.41** | 30.24% | 233.89 | 143.98 | 34.41% | 229.31 |
| 8 | 286.15 | 52.25% | 430.82 | 275.72 | **48.64%** | **409.70** | 279.33 | 51.82% | 424.89 | **271.46** | 48.97% | 428.42 | 272.45 | 50.21% | 419.20 |
| 12 | 385.61 | 68.16% | 563.90 | 380.10 | 67.51% | 546.64 | 391.35 | 69.36% | 587.54 | 382.19 | 66.10% | 578.15 | 380.73 | 64.88% | 573.84 |

When including covariates, the LSTM-SARIMAX ensemble performed better than the individual LSTM and SARIMAX models in short to medium forecast horizons. For example, for 1-week ahead forecasts, the LSTM-SARIMAX ensemble achieved the lowest MAE of 65.24 cases and a MAPE of 15.82%, outperforming the individual LSTM and SARIMAX models. The ensemble also showed the lowest RMSE of 103.52, indicating a significant improvement in predictive accuracy. However, as the forecast horizon extends, the performance advantage of the ensemble models becomes less pronounced. For instance, at 12 weeks ahead, the complex ensemble of LSTM-SARIMAX-Lag-VAR-Lag demonstrated an MAE of 380.73 cases and a MAPE of 64.88%, which was not significantly better than the individual models. This suggests that while complex ensembles may show occasional improvements, these are not consistent across all metrics, hinting at potential overfitting or inefficiencies due to increased model complexity.

Overall, the ensemble approaches show promise, particularly in short to medium-term forecasts. By combining the strengths of multiple models, the ensembles can provide more accurate and reliable predictions. However, careful consideration must be given to the potential drawbacks of increased complexity and the risk of overfitting, especially in longer forecast horizons.

### Uncertainty intervals

In this section, we consider the 95% uncertainty intervals for each of the individual models and the top performing ensemble approaches, and provide their corresponding 95% coverage probabilities and average interval widths. Table 7 and Table 9 present the 95% coverage probabilities for models using dengue cases alone and including climate covariates, respectively. Table 8 and Table 10 display the average width of the uncertainty intervals for models using dengue cases alone and including climate covariates, respectively.

**Table 7.**
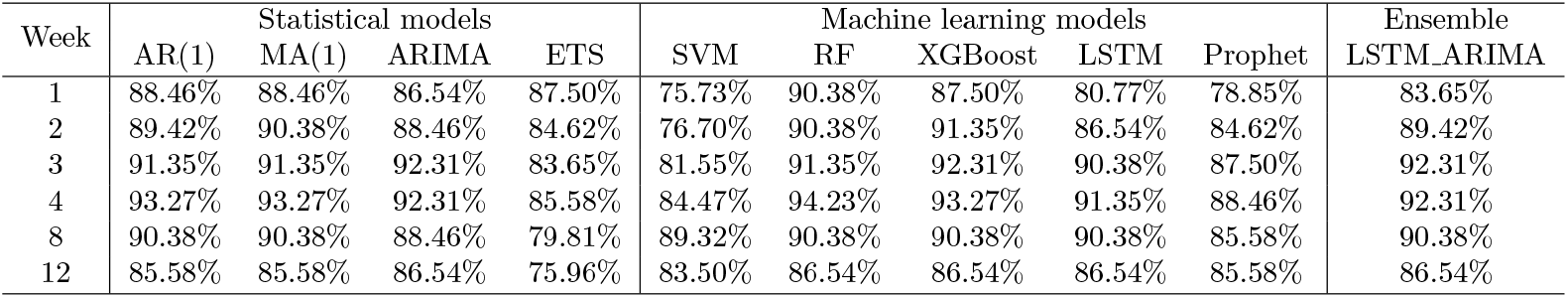
95% coverage probability of the uncertainty intervals of models using cases alone.

| Week | Statistical models |  |  |  | Machine learning models |  |  |  |  | Ensemble<br>LSTM.ARIMA |
| --- | --- | --- | --- | --- | --- | --- | --- | --- | --- | --- |
|  | AR(1) | MA(1) | ARIMA | ETS | SVM | RF | XGBoost | LSTM | Prophet |  |
| 1 | 88.46% | 88.46% | 86.54% | 87.50% | 75.73% | 90.38% | 87.50% | 80.77% | 78.85% | 83.65% |
| 2 | 89.42% | 90.38% | 88.46% | 84.62% | 76.70% | 90.38% | 91.35% | 86.54% | 84.62% | 89.42% |
| 3 | 91.35% | 91.35% | 92.31% | 83.65% | 81.55% | 91.35% | 92.31% | 90.38% | 87.50% | 92.31% |
| 4 | 93.27% | 93.27% | 92.31% | 85.58% | 84.47% | 94.23% | 93.27% | 91.35% | 88.46% | 92.31% |
| 8 | 90.38% | 90.38% | 88.46% | 79.81% | 89.32% | 90.38% | 90.38% | 90.38% | 85.58% | 90.38% |
| 12 | 85.58% | 85.58% | 86.54% | 75.96% | 83.50% | 86.54% | 86.54% | 86.54% | 85.58% | 86.54% |

**Table 8.** Average width of the uncertainty intervals of models using cases alone.

| Week | Statistical models |  |  |  | Machine learning models |  |  |  |  | Ensemble<br>LSTM.ARIMA |
| --- | --- | --- | --- | --- | --- | --- | --- | --- | --- | --- |
|  | AR(1) | MA(1) | ARIMA | ETS | SVM | RF | XGBoost | LSTM | Prophet |  |
| 1 | 362.85 | 356.35 | 395.18 | 335.26 | 412.05 | 411.82 | 478.14 | 343.56 | 858.24 | 333.64 |
| 2 | 575.24 | 563.12 | 575.84 | 480.79 | 568.55 | 615.88 | 696.32 | 514.02 | 965.86 | 468.90 |
| 3 | 716.57 | 757.53 | 741.74 | 603.25 | 712.56 | 796.76 | 903.85 | 683.67 | 1038.23 | 661.30 |
| 4 | 897.53 | 935.67 | 970.22 | 803.09 | 856.58 | 977.07 | 1045.36 | 852.74 | 1094.66 | 841.25 |
| 8 | 1440.92 | 1486.77 | 1505.23 | 1220.31 | 1353.92 | 1511.77 | 1548.26 | 1465.60 | 1346.44 | 1483.38 |
| 12 | 1881.42 | 1877.10 | 1973.47 | 1880.60 | 1678.33 | 1856.88 | 1901.31 | 1837.23 | 1579.96 | 1881.55 |

**Table 9.** 95% coverage probability of the uncertainty intervals of models including covariates.

| Week | Statistical models |  |  |  | Machine learning models |  |  |  |  | Ensemble<br>LSTM.SARIMAX |
| --- | --- | --- | --- | --- | --- | --- | --- | --- | --- | --- |
|  | SARIMAX | SARIMAX.Lag | VAR | Var.Lag | SVM | RF | XGBoost | LSTM | Prophet |  |
| 1 | 83.65% | 85.58% | 78.85% | 75.96% | 73.08% | 75.00% | 75.00% | 80.77% | 80.77% | 82.69% |
| 2 | 90.38% | 89.42% | 87.50% | 75.96% | 77.88% | 87.50% | 87.50% | 83.65% | 90.38% | 91.35% |
| 3 | 92.31% | 88.46% | 86.54% | 74.04% | 82.69% | 89.42% | 88.46% | 91.35% | 91.35% | 92.31% |
| 4 | 91.35% | 90.38% | 86.54% | 76.92% | 83.65% | 90.38% | 89.42% | 92.31% | 91.35% | 92.31% |
| 8 | 88.46% | 89.42% | 87.50% | 75.96% | 89.42% | 91.35% | 90.38% | 90.38% | 84.62% | 90.38% |
| 12 | 86.54% | 86.54% | 83.65% | 75.00% | 86.54% | 86.54% | 86.54% | 86.54% | 79.81% | 86.54% |

**Table 10.** Average width of the uncertainty intervals of models including covariates.

| Week | Statistical models |  |  |  | Machine learning models |  |  |  |  | Ensemble<br>LSTM.SARIMAX |
| --- | --- | --- | --- | --- | --- | --- | --- | --- | --- | --- |
|  | SARIMAX | SARIMAX.Lag | VAR | Var.Lag | SVM | RF | XGBoost | LSTM | Prophet |  |
| 1 | 392.05 | 412.44 | 402.72 | 354.21 | 560.02 | 1678.31 | 1668.07 | 306.98 | 795.44 | 303.93 |
| 2 | 570.76 | 531.70 | 585.18 | 533.60 | 882.41 | 1724.57 | 1723.47 | 457.93 | 918.22 | 448.20 |
| 3 | 747.85 | 679.51 | 736.93 | 710.49 | 1018.40 | 1761.29 | 1754.48 | 617.84 | 1045.29 | 639.15 |
| 4 | 964.53 | 889.74 | 945.25 | 828.14 | 1114.13 | 1812.38 | 1792.54 | 778.38 | 1172.60 | 802.21 |
| 8 | 1517.52 | 1456.77 | 1418.95 | 1421.72 | 1311.92 | 1974.08 | 1937.82 | 1368.89 | 1358.01 | 1430.66 |
| 12 | 1983.28 | 1826.20 | 1759.12 | 1708.77 | 1594.97 | 2088.01 | 2051.51 | 1761.05 | 1368.09 | 1845.90 |

As we can see from the results, all models exhibit an increase in average interval width over time, while their coverage probability remains relatively stable, near 90%. In the Supporting information, an illustration of the uncertainty intervals computed with SARIMAX and LSTM with covariates for various forecast horizons are shown in Figures S6 and S7. When using dengue case data alone, all models show consistent coverage probabilities across different forecast horizons. ARIMA, for instance, maintains a high coverage probability, averaging around 86.5% in predicting 1 week ahead and slightly increasing to 92.31% in predicting 3 weeks and 4 weeks ahead, then remain the same 86.54% at 12 weeks ahead. However, it shows one of the highest interval widths of 395.18 for 1 week ahead predictions, indicating some uncertainty in its predictions. The LSTM model, on the other hand, starts with a coverage probability of 80.77% for 1 week ahead, increasing to 86.54% by 12 weeks ahead. Its interval width is 343.56 for 1 week ahead, expanding to 1837.23 for 12 weeks ahead. Compared to ARIMA, LSTM demonstrates a slightly lower coverage probability but a more significant increase in interval width over time.

With the inclusion of climate covariates, we see improvements in the models’ performance. SARIMAX starts with a high coverage probability of 83.65% in 1 week ahead, which increases to 86.54% by 12 weeks ahead. Its interval width ranges from 392.05 in 1 week to 1983.28 in 12 weeks. Compared to ARIMA, SARIMAX shows slightly worse coverage probability but similar trends in interval width expansion. The LSTM model with climate covariates also starts strong with 80.77% coverage in 1 week and increases to 86.54% by 12 weeks. Its interval width ranges from 306.98 in week 1 to 1761.05 in week 12, showing an overall better performance than using dengue cases alone.

The ensemble models LSTM ARIMA and LSTM SARIMAX achieve a similar performance as the individual models across different weeks. The coverage probabilities are in the range of 82.69% to 92.31%. For LSTM ARIMA, the interval width starts at 333.64 in week 1 and rises to 1881.55 by week 12. LSTM SARIMAX has interval width ranging from 303.93 in 1 week to 1845.90 in 12 weeks.

### Computational efficiency

In this section, we compare the computational efficiency of the statistical models and machine learning techniques used for dengue forecasting. The computation times reported are the total times taken to generate all weekly predictions from January 2022 to December 2023 across all forecast horizons, namely, 1, 2, 3, 4, 8, and 12 weeks. The models were run on a MacBook Pro (13-inch, 2020) with processor 2 GHz quad-core Intel Core i5, and memory 16 GB 3733 MHz LPDDR4X.

Figure 5 illustrates the computational time required for running each model. The left-hand side of the bar plot displays the computational times for models using only dengue cases. On the right-hand side, the bar plot shows the computational times for models incorporating temperature and humidity covariates. Computational times are on a square root scale to accommodate the order of magnitude differences.

**Fig 5.**
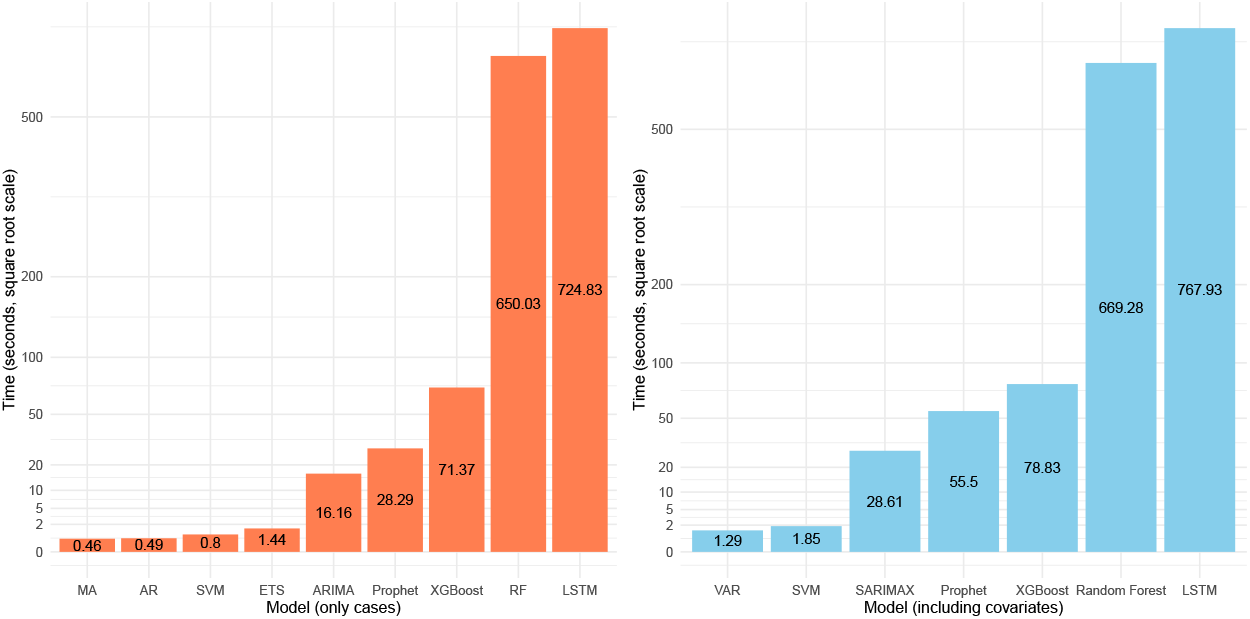
Computational time of each forecasting method.

For models using dengue cases alone, computational times vary significantly between statistical models and machine learning techniques. Statistical models, such as AR and MA, are exceptionally fast, taking less than 0.5 seconds each. This efficiency is primarily due to their simpler mathematical foundations, which involve fewer computations. ARIMA takes 16.16 seconds, which is significantly longer but still considerably faster compared to most machine learning models. The increased time for ARIMA is due to its more complex structure, which is capable of capturing intricate temporal dependencies.

In the realm of machine learning, LSTM, despite its superior predictive accuracy, has a notably high computation time of 724.83 seconds. This significant computational demand reflects the complexity of LSTM in modeling long-term dependencies and capturing intricate patterns in the data. Random Forest, another effective machine learning model, requires 650.03 seconds, also indicating its intensive computational nature. Prophet, with a computation time of 28.29 seconds, offers a more efficient alternative among machine learning models, providing a reasonable trade-off between computational cost and accuracy.

When incorporating temperature and humidity covariates, the computational times reflect similar trends. The SARIMAX model, which was highly effective in utilizing covariates for accurate predictions, takes 28.61 seconds. This is relatively efficient, considering the model’s ability to handle seasonality and multiple covariates. The VAR model, although simpler, is extremely fast at 1.29 seconds, making it suitable for quick, real-time forecasting.

Among the machine learning models, LSTM with covariates continues to require substantial computation time, clocking in at 767.93 seconds. This underscores the model’s complexity and its ability to integrate and learn from covariate data, enhancing predictive accuracy at a high computational cost. Prophet, taking 55.50 seconds, balances complexity and efficiency, making it a practical choice for many forecasting scenarios involving covariates.

In summary, statistical models like ARIMA and SARIMAX offer a good balance of computational efficiency and predictive accuracy, especially when covariates are involved. Machine learning techniques like LSTM, while providing superior accuracy, require significantly more computational resources. The choice between these models depends on the specific requirements of the forecasting task, with simpler statistical models being preferable for real-time applications in settings where computational resources are limited, and more complex machine learning models suitable for scenarios where predictive accuracy is paramount.

## Discussion

This research assesses the predictive performance and computational efficiency of a number of statistical and machine learning techniques for dengue forecasting, both with and without the inclusion of climate covariates. The study utilizes dengue cases as well as temperature and humidity in Rio de Janeiro, Brazil, a region prone to dengue outbreaks where data is readily available from the InfoDengue system [22]. The statistical models considered include AR(1), MA(1), ARIMA, ETS, VAR, and SARIMAX. Machine learning techniques utilized are SVM, Random Forest, XGBoost, LSTM, and Prophet. The study provides a thorough assessment of the forecasting performance of these methods, as well as ensemble approaches that combine individual methods, across various time frames.

Unlike other performance evaluations, we generate weekly predictions that assess the predictive accuracy of the methods at actionable scales. The flexibility in handling different forecast horizons (i.e., 1, 2, 3, 4, 8, and 12 weeks ahead) conveys the utility of the forecasting system for both immediate response planning and longer-term strategic interventions. Additionally, we compute uncertainty intervals to convey the reliability of point estimates. Our evaluation also includes the computational efficiency of each model, which is an important consideration in resource-constrained environments.

In our implementation, we use a moving window strategy that allows models to continuously adapt to new data, capturing the evolving patterns and trends in dengue incidence. This adaptability is crucial for accurate predictions, especially in the context of a disease influenced by various fluctuating factors such as climate, population movement, and public health interventions. The results highlight the nuanced capabilities of each method, which can inform the implementation of dengue surveillance systems and the allocation of resources to combat dengue outbreaks.

Among the statistical models evaluated, ARIMA emerged as the best model when using only historical case data. Its simplicity, rapidity and robust predictive accuracy make it a reliable choice for short to medium-term forecasts. ARIMA’s ability to capture temporal dependencies effectively contributed to its strong performance. However, the inclusion of climate covariates significantly enhanced its predictive power through the SARIMAX model. By accounting for climate factors such as temperature and humidity, SARIMAX provided a more comprehensive analysis, leading to improved accuracy. Moreover, the use of lagged covariates in SARIMAX further enhanced long-term prediction capabilities, addressing the inherent uncertainties associated with extended forecast horizons.

The Long Short-Term Memory (LSTM) model, particularly when combined with climate covariates, proved to be the most accurate machine learning model overall. LSTM’s recurrent neural network structure excels at handling non-linear temporal patterns, making it highly effective in capturing the complex dynamics influenced by climate factors. This resulted in consistently lower errors across all forecast horizons. Despite being slower to train and predict due to its computational complexity, LSTM’s superior accuracy makes it an unmatched choice for highly precise predictions. The model’s ability to integrate and learn from additional covariate data further bolstered its performance, especially in the context of medium to long-term forecasts.

For long-term forecasts (i.e., 12 weeks), the Prophet model with climate covariates demonstrated the best accuracy. Prophet’s strength lies in its ability to identify and adapt to long-term patterns, making it particularly effective for distant predictions. The inclusion of climate covariates enabled Prophet to capture seasonal variations and other long-term trends more accurately. While Prophet may not match the short-term accuracy of models like LSTM or ARIMA, its performance in long-term forecasting highlights its utility in scenarios where understanding and predicting extended trends are crucial.

In addition to evaluating individual models, we explored the potential benefits of ensemble approaches by combining the strengths of the best-performing statistical and machine learning models. By averaging forecasts from both statistical and machine learning models, the ensembles capitalized on the strengths of each method, resulting in more robust and reliable predictions. The use of ensemble models, particularly those combining ARIMA and LSTM for cases alone and SARIMAX with LSTM for models including covariates, demonstrates significant promise in advancing the accuracy and reliability of dengue predictions.

Our study has some limitations that indicate areas for future research. First, we evaluate the methods in a single geographical location, namely, Rio de Janeiro. Future research could explore the generalizability of these models by applying them to different geographical areas with different climatic and socio-economic conditions. This would help to validate the models’ robustness and enhance their utility in diverse settings. In addition, we considered temperature and humidity as covariates in some of the models. While these are critical factors for dengue transmission, additional variables such as population density, mobility patterns, socio-economic factors, and land use changes could also play significant roles in dengue dynamics [11]. Future studies should consider incorporating a broader range of predictive factors which could improve the models’ accuracy and provide a more comprehensive understanding of dengue transmission mechanisms.

Moreover, we primarily focused on the assessment of the predictive accuracy and computational efficiency of different models. While these are crucial aspects, future research could also examine the interpretability and usability of the models from a public health perspective. Ensuring that the models are not only accurate but also interpretable is essential to implement strategies for disease prevention and control. Lastly, exploring the use of spatial models that borrow information of close or connected regions [51, 52], and advanced machine learning techniques, such as deep reinforcement learning [53, 54], could further enhance dengue forecasting capabilities.

To conclude, this study provides a thorough evaluation of dengue prediction methods, showcasing how statistical methods, advanced machine learning models, and climate covariates yield valuable insights for proactive disease surveillance. Our findings provide evidence to inform the development of more robust and comprehensive models that better support public health efforts. By leveraging dengue forecasts, officials can optimize resource allocation, implement timely control measures, and reduce the impact of dengue outbreaks on the population.

## Data Availability

All relevant data are within the manuscript and its Supporting Information files.

## Supporting information

**S1 Comparison of the predictions obtained by the best performing statistical and machine learning techniques for different time horizons**.

**S2 Boxplots of the absolute errors obtained by the forecasting methods when using only cases**.

**S3 Boxplots of the absolute percentage errors obtained by the forecasting methods when using only cases**.

**S4 Boxplots of the absolute errors obtained by the forecasting methods when including covariates**. Ensemble refers to the best ensemble approach using covariates which is LSTM and SARIMAX.

**S5 Boxplots of the absolute percentage errors obtained by the forecasting methods including covariates**. Ensemble refers to the best ensemble approach using covariates which is LSTM and SARIMAX.

**S6 Real cases, predictions, and 95% uncertainty intervals computed with SARIMAX across various forecast horizons**.

**S7 Real cases, predictions, and 95% uncertainty intervals computed with LSTM including covariates across various forecast horizons**.

